# Ocrelizumab versus Natalizumab in Relapsing-Remitting Multiple Sclerosis: A Registry-Linked Electronic Health Records Study

**DOI:** 10.64898/2025.12.01.25341331

**Authors:** Feiqing Huang, Wen Zhu, Jue Hou, Sara Morini Sweet, Yunqing Han, Jun Wen, Katherine P. Liao, Tianrun Cai, Tanuja Chitnis, Florence T. Bourgeois, Zongqi Xia, Tianxi Cai

**Affiliations:** Department of Biostatistics, Harvard T.H. Chan School of Public Health, Boston, MA, USA; Division of Biostatistics and Health Data Science, School of Public Health, University of Minnesota, Minneapolis, MN, USA; Harvard-MIT Center for Regulatory Science, Harvard Medical School, Boston, MA, USA; Pediatric Therapeutics and Regulatory Science Initiative, Computational Health Informatics Program (CHIP), Boston Children’s Hospital, Boston, MA, USA; Department of Biomedical Informatics, Harvard Medical School, Boston, MA, USA; Department of Neurology, University of Pittsburgh, Pittsburgh, PA, USA; Division of Rheumatology, Inflammation, and Immunity, Mass General Brigham, Boston, MA, USA; Department of Neurology, Mass General Brigham, Boston, MA, USA

**Author notes:** **Corresponding Authors**: Tianxi Cai, ScD, Department of Biomedical Informatics, Harvard Medical School, Address: 10 Shattuck Street, Room 434, Boston, MA 02115, Zongqi Xia, MD, PhD, Department of Neurology, University of Pittsburgh, Address: 3501 Fifth Ave, BST3-10.044.

**Keywords:** real-world evidence, electronic health records, imputation, confounding, multiple sclerosis, disability

## Abstract

**Background:** Ocrelizumab and natalizumab are commonly prescribed high-effectiveness disease-modifying therapies (DMTs) for relapsing-remitting multiple sclerosis (RRMS). However, no randomized clinical trial and few real-world studies have directly compared their effectiveness in reducing disability progression. Subtype classification and disability status are critical for multiple sclerosis (MS) research, but these data are often missing in electronic health records (EHRs), limiting robust real-world evidence generation.

**Objective:** To compare the effectiveness of ocrelizumab and natalizumab in two-year clinician-rated disability progression among RRMS patients using longitudinal registry-linked EHR data.

**Design:** Retrospective cohort study.

**Setting:** A large healthcare system that includes both academic and community practices.

**Participants:** Patients diagnosed with MS who initiated ocrelizumab or natalizumab between 2012 and 2020, with at least 6-month EHR data before treatment initiation and no prior exposure to other high-effectiveness DMTs.

**Exposures:** Treatment with ocrelizumab vs natalizumab.

**Measurements:** We developed an ensemble machine learning model to impute RRMS subtype and disability outcomes using structured and narrative EHR data. The primary outcome was moderate/severe clinician-rated disability at 2 years (observed or imputed Expanded Disability Status Scale [EDSS]≥4) after treatment initiation. We estimated the average treatment effects using semi-supervised doubly robust approach with comprehensive confounder adjustment and calibration to mitigate imputation bias. Covariates included standard demographic and clinical features such as baseline disability as well as knowledge graph-selected features. Sensitivity analyses used observed EDSS scores in registry-derived RRMS patients. Exploratory analyses included rituximab, another B-cell–depleting therapy, with adjustments for differences in patient profiles.

**Results:** Among RRMS patients, those treated with ocrelizumab (n=543) had a significantly lower two-year risk of moderate/severe disability compared with those treated with natalizumab (n=205) based on imputed outcomes (risk difference, –5.87%; 95% CI: –11.28% to –0.46%; p=0.033) after confounder adjustment. Sensitivity analyses yielded consistent findings using imputed or observed EDSS outcomes in registry-derived RRMS patients.

**Conclusion and relevance:** In this real-world comparative effectiveness study using a novel semi-supervised doubly-robust framework, ocrelizumab was associated with a lower risk of disability progression than natalizumab among RRMS patients. This approach provides a roadmap for generating robust large-scale real-world evidence in settings of missing key inclusion features and outcomes.

## INTRODUCTION

Multiple sclerosis (MS) is a chronic autoimmune disease that causes inflammatory demyelination in the central nervous system and progressive neurodegeneration, leading to disability.^1^ Among more than 20 approved MS disease-modifying therapies (DMTs) with distinct mechanisms of action, robust comparative evidence for therapies of similar perceived effectiveness remains lacking. Within higher-effectiveness options, natalizumab [NTZ] and ocrelizumab [OCR] are widely prescribed for relapsing-remitting MS (RRMS) and associated with reduced relapse.^2–9^ However, no randomized controlled trial (RCT) and few real-world studies have compared their long-term effects on disability.^5–10^

RRMS is more common than primary progressive MS (PPMS) and secondary progressive MS (SPMS) subtype.^11^ MS subtype classification informs RCT eligibility, confounder adjustment in observational studies, and treatment decisions in clinical practice. While research registries often record MS subtype, it is rarely captured in electronic health records (EHR).^12^

The Expanded Disability Status Scale (EDSS) is a clinician-rated measure of neurological impairment across eight functional systems. Higher EDSS scores indicate greater disability.^13,14^ Although EDSS is frequently used to evaluate disability outcome in RCTs, it has limitations. Like MS subtype, EDSS scores are typically documented in registries but not in EHRs.

Integrating registry and EHR data can strengthen real-world evidence by combining disease subtype and outcome measures (often not recorded during routine care) from registries with longitudinal clinical data besides the disease of interest (rarely recorded in disease registries) from EHR. Here, we present a framework for linking registry and EHR data to phenotype MS subtype and EDSS outcomes beyond registry populations. As a clinically relevant illustration, we compared the effectiveness of OCR versus NTZ in delaying two-year disability progression among patients with imputed or registry-derived RRMS subtype.

## METHODS

### Ethics Approval

The institutional review boards of the University of Pittsburgh (STUDY20070274) and Mass General Brigham (2023P001450) approved the study protocols. The use of de-identified clinical data was deemed exempt.

### Data Sources and Patient Cohort

Figure 1 illustrated the overall study design. This study leveraged registry-linked EHR data.^15–19^ EHR data from Mass General Brigham (MGB) included both academic and community practices from January 1, 2000, to December 31, 2023. The Comprehensive Longitudinal Investigation of Multiple Sclerosis at Brigham and Women’s Hospital (CLIMB) registry is a prospective cohort that has followed MS patients with standardized clinical visits, including EDSS assessments, since 2000.^20^ To construct the EHR-based MS cohort, we first identified all patients with at least one MS diagnosis code. Given the high sensitivity but low specificity of ICD codes for disease classification,^21^we then applied a knowledge graph-guided “weakly supervised” phenotyping algorithm^22^ to improve diagnostic accuracy and reduce misclassification. Using a classification threshold calibrated to achieve 90% specificity (reaching 92% positive predictive value, PPV) through validation against a manually annotated subset of MS diagnosis, we identified 18,560 patients for downstream imputation of RRMS subtype and EDSS and causal analysis.^19^

**Figure 1.**
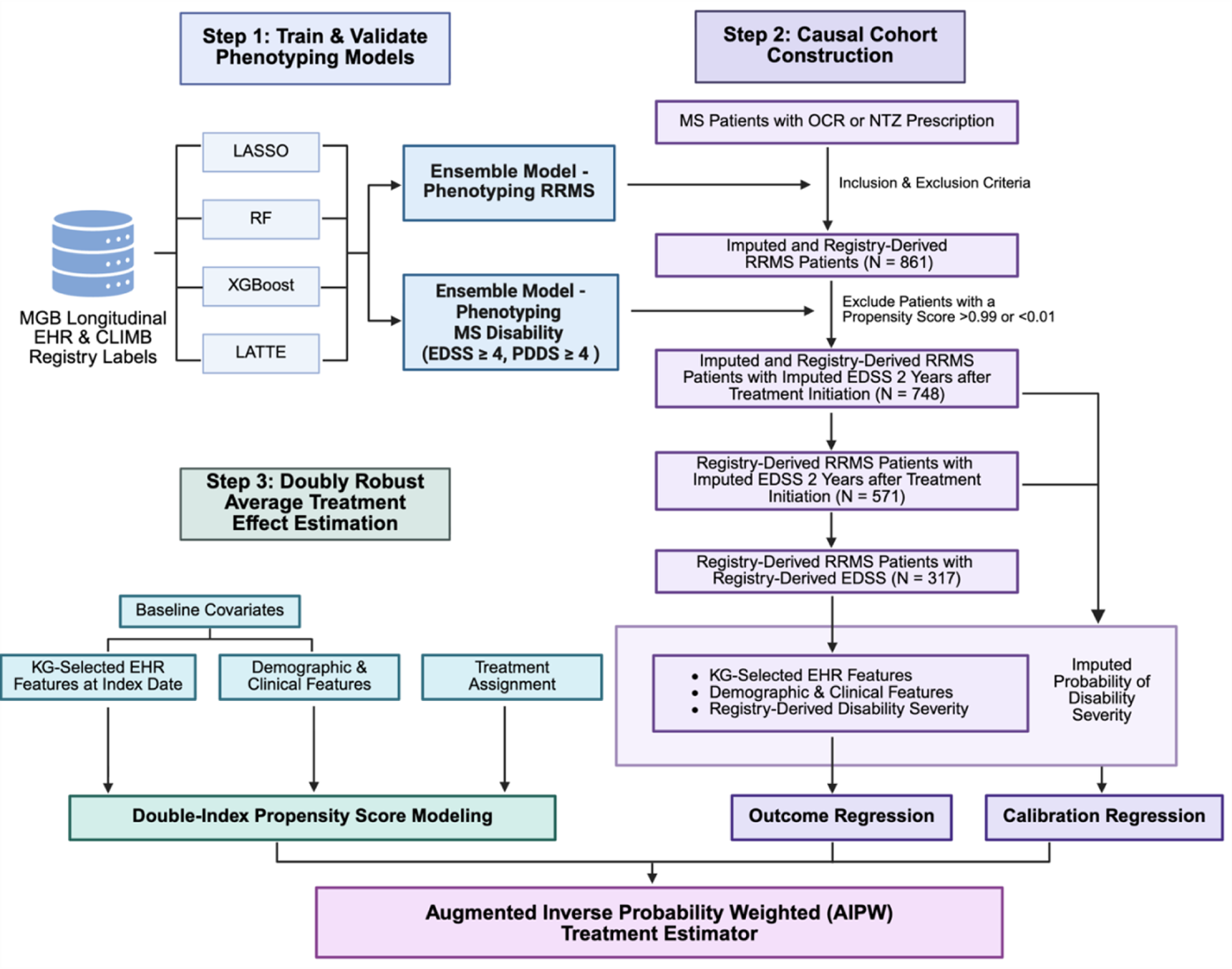
Overview of the study design and the patient cohort for causal inference. *EDSS*, Expanded Disability Status Scale; *EHR*, electronic health records; *KG*, knowledge graph; *LASSO*, Least Absolute Shrinkage and Selection Operator; *LATTE*, Label-efficient Incident phenoTyping; *NTZ*, natalizumab; *OCR*, ocrelizumab; *RF*, random forest; *RRMS*, relapsing-remitting multiple sclerosis; *XGBoost*, extreme gradient boosting.

### Feature Set for Imputation and Confounder Adjustment

We included EHR features to achieve two objectives: (1) impute RRMS subtype and disability outcome, and (2) adjust for potential confounding in causal analyses.

For each patient, we extracted monthly counts of codified EHR data, including diagnosis codes (*e.g.*, International Classification of Disease [ICD]), procedure codes (*e.g.*, Current Procedural Terminology [CPT]), prescriptions (*e.g.*, RxNorm), and laboratory tests (*e.g.*, Logical Observation Identifiers Names and Codes [LOINC]).^23^ We mapped all ICD codes to PheCodes^21^, consolidated CPT codes using the Clinical Classifications Software (CCS) for Services and Procedures, and grouped electronic prescription codes at the RxNorm ingredient level.^24^ Clinical notes were processed using a natural language processing (NLP)^25,26^ tool to generate monthly counts of medical terms, which were mapped to Concept Unique Identifiers (CUIs) according to the Unified Medical Language System (UMLS).^27^ MS-relevant codes and medical terms were identified using multi-source clinical knowledge graphs.^22,28^ To account for healthcare utilization, we quantified the total counts of codified EHR features. Counts of codified and narrative EHR features were log transformed.

To further adjust for confounding, we included standard demographic and clinical features commonly used in RCTs: *i.e.,* age at treatment initiation, sex (male vs female), race and ethnicity (non-Hispanic White vs otherwise), disease duration (defined as the time from the first recorded MS PheCode to treatment initiation, *i.e.,* index date) and prior MS-related medications.

Informative covariates were shown in **Supplementary Table 1**.

### RRMS Subtype Imputation

MS subtypes in the CLIMB registry were recorded longitudinally at irregular intervals (*i.e.,* a median of three entries per patient). For patients with linked EHR data, registry-derived RRMS based on clinician documentation were used as the gold-standard RRMS status (*i.e.,* observed RRMS) at the corresponding time for training the subtype imputation model.

To account for changes in MS subtypes, models were trained to impute RRMS status (*i.e.,* imputed RRMS) using features (see Method - Feature Set) aggregated over consecutive two-year windows. RRMS classification models were developed using both standard supervised learning methods (*i.e.,* LASSO, XGBoost, Random Forest) and a semi-supervised approach (*i.e.,* LATTE, Label-efficient incident phenotyping)^29^, which additionally leveraged surrogates and EHR data from patients outside the registry. Final RRMS classifications were derived using a cross-validated ensemble of all models (*i.e.,* “Ensemble” method).

Imputation model performance was evaluated in the held-out set comprising 25% of registry patients excluded from training. For the RRMS subtype classification, we reported the area under the receiver operating characteristic curve (AUC), sensitivity, specificity, and PPV.

### Clinician-Rated Disability Outcome Imputation

The clinician-rated disability measure based on EDSS ranges from 0 to 10 in 0.5 increment. As EDSS of 4.0 typically represents disability threshold significant enough to impact patient life, we operationally classified EDSS ≥4.0 as “moderate/severe disability,” and <4.0 as “no/mild disability”. Given that EDSS scores in the CLIMB registry were typically recorded every six months (*i.e.,* a median of four assessments per patient), we computed the median EDSS score and defined binary threshold (observed EDSS ≥4.0 vs <4.0) within consecutive six-month windows as the gold-standard clinician-rated disability labels.

To account for changes in EDSS scores and impute missing scores, models were trained to impute binary EDSS threshold (*i.e.,* imputed EDSS) using features (see Method - Feature Set) aggregated over consecutive 6-month windows. We trained models using data from a randomly selected subset of 500 patients with available EDSS scores. Binary EDSS classification models were developed using standard supervised learning methods (*i.e.,* LASSO, XGBoost, Random Forest) and the semi-supervised LATTE method. Predicted likelihood (*i.e.,* probability) for moderate or severe clinician-rated disability was derived using a cross-validated ensemble of all models (*i.e.,* “Ensemble” method).

Imputation model performance was evaluated in the held-out set comprising registry patients excluded from training by comparing predicted likelihood of moderate/severe disability to the gold-standard EDSS≥4.0 labels. For the binary EDSS classification, we reported AUC, sensitivity, specificity, and PPV.

### Patient-Reported Disability Outcome Imputation

The patient-reported disability measure based on Patient Determined Disease Steps (PDDS) provides crucial patient perspectives and complements clinician-rated EDSS score.^30^ Thus, we also imputed baseline PDDS scores (See methods in **Supplementary Materials S1.1)**.

### DMT Choice and Target Population

To illustrate a framework for scalable and robust RWE generation, we set out to compare two DMTs that are both considered highly effective for MS but lack head-to-head RCT evidence. Only one DMT (*i.e.,* OCR) is approved for PPMS and one DMT (*i.e.,* mitoxantrone, rarely prescribed) has been approved for non-active SPMS to date. In contrast, multiple DMTs are approved for RRMS. Thus, we focused on two commonly prescribed DMTs for RRMS: OCR and NTZ.

The target population in this study were patients with the RRMS subtype at treatment initiation. Baseline RRMS status was determined using either registry-derived labels or imputed labels, with the latter classified using a threshold calibrated to achieve PPV≥0.90.

Inclusion criteria included: (1) treatment initialization between 2012 and 2020 to ensure sufficient pre-treatment and post-treatment follow-up, (2) no prior exposure to other high-effectiveness DMTs or chemotherapy agents (*e.g.*, Alemtuzumab, Cyclophosphamide, Cladribine, Mitoxantrone and Rituximab [RTX]), and (3) at least six months of available EHR data before and after the index date (see Treatment Exposure).

### Treatment Exposure

For the *primary* causal analysis, we compared OCR and NTZ. For the *exploratory* causal analysis, we compared B-cell depletion therapy (including OCR and RTX) and NTZ (**Supplementary Material S1.2, Supplementary Figure 1**). Treatment initiation was defined as the first recorded instance of the corresponding medication code in the EHR and designated as the index date (*i.e.,* baseline). Patients were assigned to the target exposure group based on the first treatment received.

### Treatment Outcome

The primary study outcome was the likelihood of developing moderate/severe disability (EDSS≥4.0) during two years after target treatment initiation, modeled as a continuous probability between 0 and 1 and denoted as pEDSS. We included observed EDSS scores when available and imputed EDSS scores when gold-standard labels are unavailable. To address temporal fluctuations in EDSS scores and noise in observed measurement or imputation, we first estimated pEDSS in each of the four consecutive 6-months periods following treatment initiation and then aggregated these estimates to derive a two-year cumulative likelihood. This approach provides a more robust representation of overall disability status during the two-year follow-up.

### Statistical Analysis for Estimating Average Treatment Effect

We estimated the average treatment effect (ATE) using a doubly robust causal inference framework that combined propensity score modeling with calibration regression on imputed outcomes to minimize bias.

To adjust for potential confounding, the analysis included 254 features measured at the index date, including (1) standard demographic and clinical features (see Feature Set): *e.g.*, age, sex, race and ethnicity, and disease duration as well as imputed likelihood of binary baseline clinician-rated and patient-reported disability status during the two years preceding the index date; (2) EHR features relevant to “multiple sclerosis” or “disability”, selected using multi-source clinical knowledge graphs ^19,22^ (*e.g.,* MS diagnosis codes, mentions of “magnetic resonance imaging” in clinical notes) and aggregated over the two years preceding the index date; (3) interaction terms between baseline disability status and other covariates to enable stratification by disability status. Covariate balance was assessed by comparing weighted covariate distributions between treatment exposure groups using tests of independence.

Propensity scores for treatment exposure and calibration regressions were estimated using penalized logistic regression, which automatically selected informative covariates while shrinking non-informative ones. We excluded patients with extreme propensity scores (≥0.99 or ≤0.01) since they were highly unlikely to have received the alternative treatment, limiting the ability to infer counterfactual outcomes. We then applied augmented inverse probability weighting (AIPW) to integrate these components and compute the ATE.

To assess robustness, we conducted two sensitivity analyses to evaluate the consistency of ATE estimates when using gold-standard labels rather than imputed ones: (1) restricting to patients with registry-derived RRMS subtype; (2) restricting to patients with both registry-derived RRMS subtype and observed EDSS assessments within the two-year follow-up window.

## RESULTS

### RRMS Subtype and Disability Status Imputation

For imputing the RRMS subtype, LATTE achieved an AUC of 0.879 in the held-out test set, while the Ensemble method combining the machine learning methods marginally enhanced the prediction to AUC of 0.894 (**Table 1**). Using a pre-specified calibrated threshold of ≥0.63 for the RRMS likelihood (to reach PPV≥0.90), the ensemble model achieved a sensitivity of 0.808, specificity of 0.831 and PPV of 0.901 and was used to define a cohort with imputed RRMS subtype (**Supplementary Table 5**).

**Table 1.**
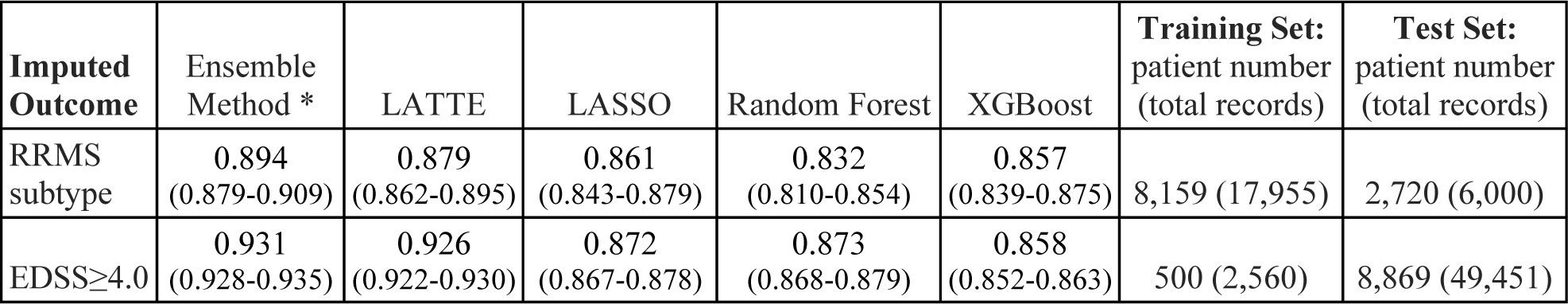
Imputation predictive performance. Area under the receiver operating characteristic curve (AUC) with 95% confidence intervals for imputing the **RRMS subtype** and for imputing clinician-rated moderate to severe disability (**EDSS≥4.0**) by the different machine learning methods is shown.

| **Imputed Outcome** | **Ensemble Method \*** | **LATTE** | **LASSO** | **Random Forest** | **XGBoost** | **Training Set:**<br>**patient number**<br>**(total records)** | **Test Set:**<br>**patient number**<br>**(total records)** |
| --- | --- | --- | --- | --- | --- | --- | --- |
| RRMS subtype | 0.894<br>(0.879-0.909) | 0.879<br>(0.862-0.895) | 0.861<br>(0.843-0.879) | 0.832<br>(0.810-0.854) | 0.857<br>(0.839-0.875) | 8,159 (17,955) | 2,720 (6,000) |
| EDSS≥4.0 | 0.931<br>(0.928-0.935) | 0.926<br>(0.922-0.930) | 0.872<br>(0.867-0.878) | 0.873<br>(0.868-0.879) | 0.858<br>(0.852-0.863) | 500 (2,560) | 8,869 (49,451) |

For imputing the clinician-rated binary disability status (*i.e.,* likelihood of EDSS≥4.0) either during pre-treatment baseline or treatment follow-up, LATTE achieved an AUC of 0.926 in the held-out test set, while the Ensemble method again slightly improved the prediction to AUC of 0.931 (**Table 1**). Using a representative calibrated threshold of 0.78 for pEDSS≥4.0 (to reach PPV≥0.90) for reporting and compared against observed labels, the ensemble model, which achieved a sensitivity of 0.506, specificity of 0.983, and PPV of 0.902 (at the representative threshold), was used for defining baseline and post-treatment clinician-rated disability (**Supplementary Table 6**). Unlike the RRMS imputation to establish a cohort with the RRMS subtype, disability imputation (pEDSS, likelihood value ranging between 0 and 1) did not require a cut-off calibration threshold.

Please see **Supplementary Material 1.2**, **Supplementary Table 2** and **Supplementary Table 7** for performance metrics of imputing the patient-reported binary disability status. Again, the ensemble method was used for defining baseline patient-reported disability.

### Patient Profile in the Causal Analysis Cohort

For the causal analysis cohort comparing OCR vs NTZ, we excluded 113 out of the 861 eligible patients with imputed RRMS subtype due to extreme propensity scores, leaving a final cohort of 748 patients with imputed RRMS subtype receiving either treatment (543 OCR vs 205 NTZ) (Figure 1). Before covariate adjustment, treatment groups differed in baseline characteristics (**Supplementary Table 1**). Compared with OCR-treated patients at baseline, NTZ-treated patients had higher predicted pre-treatment disability measured by EDSS (OCR: 0.16 vs NTZ: 0.21; *p*=0.003), higher predicted pre-treatment disability measured by PDDS (OCR: 0.12 vs NTZ: 0.14; *p*=0.02), lower healthcare usage measured by EHR codes (log-counts OCR: 2.74 vs NTZ: 2.40; *p*<0.001) and fewer MS diagnosis codes (log-counts OCR: 2.41 vs NTZ: 2.03, *p*<0.001). These baseline differences suggest potential confounding by disease severity and healthcare utilization. After covariate adjustment by IPW, treatment groups were balanced on baseline characteristics (Figure 2).

**Figure 2.**
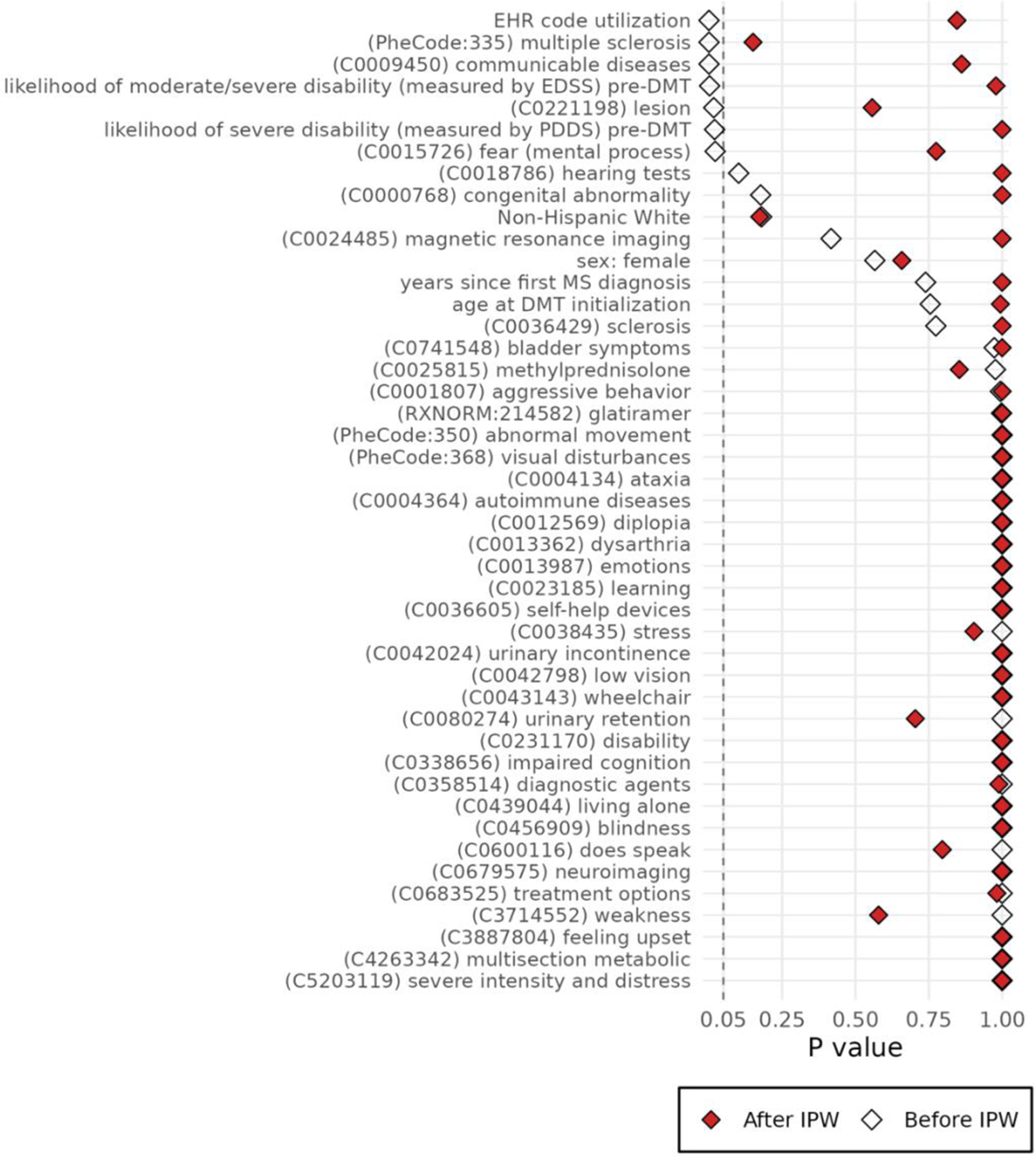
Covariate balancing. P-values before and after inverse probability weighting (IPW) are shown for all covariates with non-zero coefficients in either the propensity score or outcome regression models in the **primary causal analysis** comparing the 2-year disability between **OCR** versus **NTZ**. The vertical dashed line at *p*=0.05 indicates the threshold for nominal significance. P-values were derived from chi-squared tests for categorical variables or two-sample Kolmogorov–Smirnov tests for continuous variables. All covariates were marginally balanced (*p*>0.05) post-weighting.

### Primary Causal Analysis

In the primary causal analysis using imputed RRMS subtype and imputed disability outcome (*i.e.,* pEDSS≥4.0 during two years after treatment initiation), OCR-treated patients had a lower two-year risk of moderate to severe disability (5.38%, 95% CI: 3.40%-7.36%) compared with NTZ-treated patients (11.25%, 95% CI: 6.79%-15.71%). The absolute risk difference was –5.87% (95% CI: –11.28% to –0.46%; *p*=0.033), favoring OCR (Figure 3).

**Figure 3.**
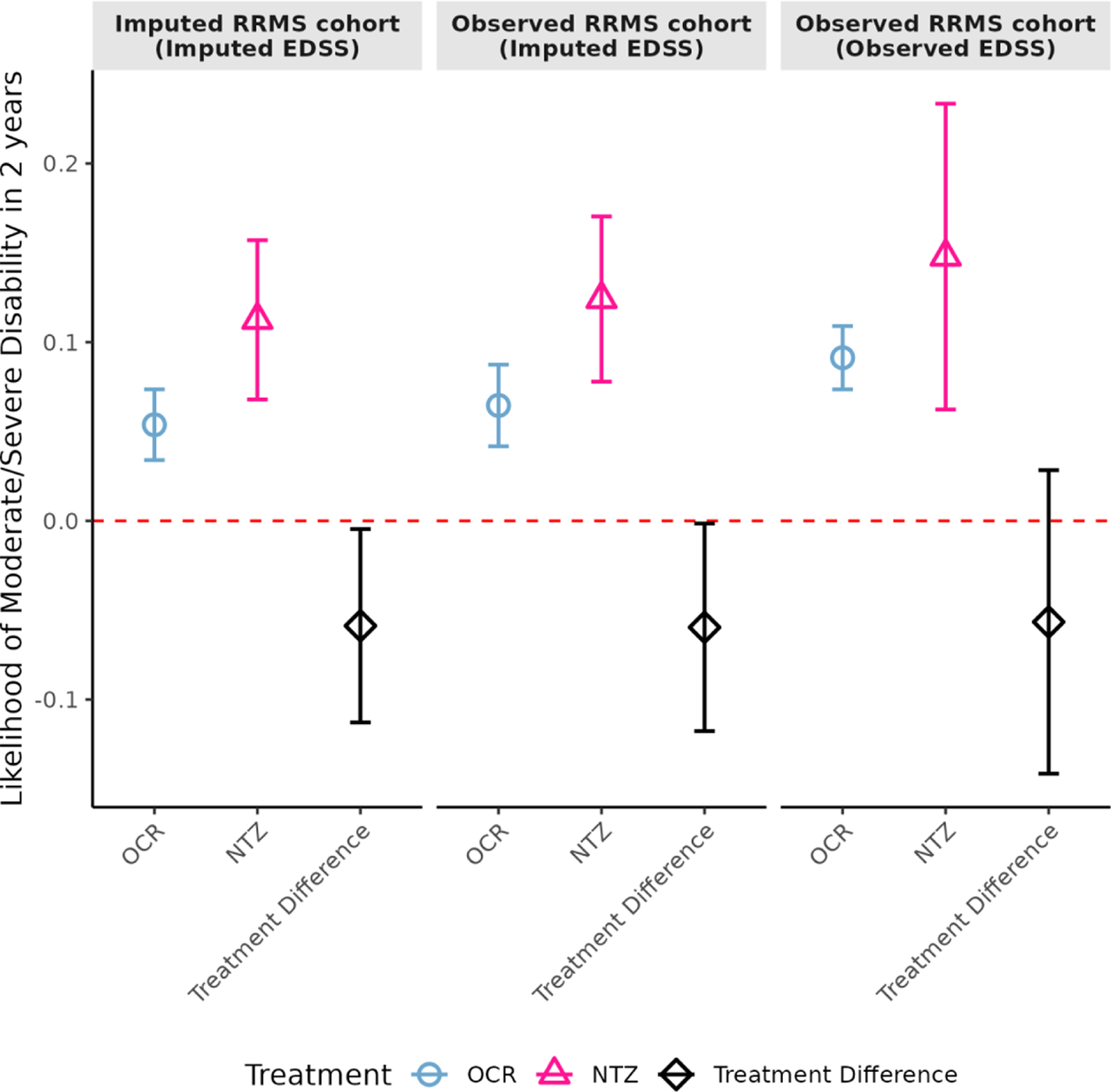
Average treatment effects of OCR vs NTZ. Estimated average treatment effects (markers) on pEDSS (*i.e.,* imputed likelihood of moderate or severe disability, EDSS≥4.0) during the two years after treatment initiation are shown with pointwise 95% confidence intervals (vertical bars). OCR was associated with a lower two-year risk of moderate or severe disability across analyses, when using imputed EDSS in patients with imputed RRMS subtype, imputed EDSS in registry patients with observed RRMS subtype or observed EDSS in registry patients with observed RRMS subtype. While point estimates remained similar, the confidence intervals widened when using registry-derived subtype and disability status.

### Sensitivity Causal Analysis

Sensitivity analyses confirmed that ATE estimates remained consistent when using gold-standard labels (Figure 3). When analyzing 571 patients with a registry-derived RRMS subtype, OCR was associated with a lower two-year disability risk (6.46%, 95% CI: 4.17%–8.74%) compared with NTZ (12.42%, 95% CI: 7.79%–17.04%), yielding a statistically significant risk difference of – 5.96% (95% CI: –11.77% to –0.15%; *p*=0.045) favoring OCR. When analyzing 317 patients with both a registry-derived RRMS subtype and observed EDSS outcomes (*i.e.,* gold-standard observed labels only), OCR also showed a lower two-year disability risk (9.13%, 95% CI: 7.36%–10.9%) compared with NTZ (14.79%, 95% CI: 6.23%–23.35%), but the difference was not statistically significant (risk difference –5.66%; 95% CI: –14.15% to 2.84%; *p*=0.192), likely reflecting the smaller sample size and greater variability.

### Exploratory Causal Analysis

During the study period, RTX was commonly used off label in RRMS. For exploratory causal analysis comparing B-Cell Depletion therapy (BCD: OCR or RTX) with NTZ, BCD was associated with lower two-year disability risk than NTZ across all population definitions (**Supplementary Materials S1.2, Supplementary Figure 2-3, Supplementary Table 3-4**.)

## DISCUSSION

This study aimed to strengthen RWE for head-to-head comparison of treatment effects on disability outcomes between OCR and NTZ in RRMS given the absence of RCT evidence. We integrated complementary registry and EHR data from a large healthcare system, applied ensemble machine learning methods to impute key missing patient information (*i.e.,* RRMS subtype, disability status), and developed a novel semi-supervised doubly robust approach with a calibration step to mitigate imputation bias. This new framework enables direct comparison of treatment effects on disability outcomes in target MS populations using large-scale real-world clinical data beyond registry patients. In the primary causal analysis as a clinically relevant illustration of the framework, OCR was associated with a significantly lower two-year risk of disability progression compared with NTZ in populations defined by both imputed and registry-derived RRMS subtype.

### Findings in Context and Clinical Implications

To our knowledge, previous observational studies comparing OCR and NTZ relied exclusively on registry data, without extending analyses to patients outside registries or integrating EHR data that could provide complementary and informative patient characteristics often missing from disease-specific registries^31–36^. While most prior studies reported no significant differences between OCR and NTZ across various definitions of disability progression, Yamout et al. observed higher rates of *disability improvement* with NTZ compared with BCD therapy (*i.e.,* OCR and RTX) in RRMS.^32^

Several key differences distinguish our study from the Yamout study beyond leveraging complementary data sources (registry-linked EHR cohort vs registry only). First, OCR- and RTX-treated patients in our study had distinct profiles, necessitating careful patient projection to the same target population before grouping both DMTs under BCD for comparison with NTZ, a step not addressed in the Yamout study. Second, the studies used different disability outcome definitions. The two-year risk of moderate or severe disability, aggregated across four consecutive 6-month windows in our study, is likely more stable than the 6-month confirmed disability change used in the Yamout study. Notably, Yamout et al. also reported no significant difference in disability progression between BCD and NTZ. Third, our doubly robust causal estimation approach is less vulnerable to model misspecification than the propensity scores matching method used in the Yamout study. Finally, differences in patient populations (United States vs Middle Eastern and North African) may further explain the divergent findings. Future RCTs are needed to provide the definitive evidence regarding whether OCR is superior to NTZ in delaying disability progression in target populations.

### Inclusion of Rituximab with Ocrelizumab for Causal Analysis

An important and perhaps unsurprising conclusion from this study is the need to project RTX- and OCR-treated patients onto a common target population for causal analysis, given the distinct patient profiles of these treatment groups. In MS research, DMTs with similar mechanisms of action are typically grouped together: *e.g.*, RTX and OCR under BCD therapy. However, their use in routine practice could differ substantially.

RTX is frequently prescribed off label, giving clinicians greater flexibility in dosing and frequency compared with OCR. In contrast, OCR administration is typically restricted by insurance requirements to a fixed regimen (*i.e.,* 600 mg every six months) according to the drug approval. Clinically, RTX is more often administered in patients with more aggressive disease because of this flexibility for higher dose and frequency.

Prior literature supports our observation of differences between RTX and OCR groups^37–42^: RTX-treated patients tend to have longer disease duration, higher baseline disability scores, and more prior DMT exposure. Prior studies also reported higher relapse rates, greater likelihood of treatment discontinuation, increased hospitalization risk, and different safety profiles for RTX compared with OCR. These differences underscore the need for caution and care when grouping RTX and OCR under BCD therapy, as selection and indication biases may confound results.

To address this in our exploratory causal analysis comparing BCD (OCR or RTX) vs NTZ, we pre-defined a common target population and then projected patients receiving different treatments onto that shared covariate space. Specifically, RTX-treated patients in the exploratory analysis were mapped to the covariate distribution of the primary causal analysis cohort (OCR vs NTZ) (see **Supplementary Material S1.2**).

Future studies could conduct separate head-to-head comparisons of RTX vs NTZ and RTX vs OCR using methods that explicitly account for differences in patient profiles. Such analyses will help clarify whether observed heterogeneity reflects treatment selection patterns or true differences in treatment effectiveness.

### Strengths

This study has several noteworthy strengths. First, by linking registry and EHR data, we expanded the target population beyond registry patients for causal analysis. Leveraging gold-standard registry labels, imputing key missing features (*i.e.,* RRMS subtype, baseline disability status, and two-year disability outcome) from EHR data using ensemble machine learning more than doubled the sample size (*i.e.,* 2.36-fold, from 317 to 748 eligible patients), increasing statistical power and improving the generalizability beyond registry-only cohorts.

Second, our causal analysis incorporated a rich set of covariates selected through multi-source clinical knowledge graphs, including NLP-extracted concepts, alongside standard clinical and demographic features, patient-reported and clinician-rated baseline disability status, and healthcare utilization, many of which were not considered in prior studies.

Third, our novel semi-supervised doubly robust causal estimation framework, which combines machine learning-based imputation with a calibration step to mitigate bias, addresses major challenges in real-world data where key patient profiles and outcomes are often incomplete. This approach ensures reliable and interpretable treatment effect estimates in routine clinical settings. Importantly, this study demonstrates the feasibility of scaling comparative effectiveness research using real-world data while maintaining methodological rigor.

Finally, it provides methodologically robust and clinically relevant real-world evidence of comparative effectiveness (*i.e.,* OCR vs NTZ in RRMS) to bridge the gap left by the absence of head-to-head RCTs, offering practical guidance for MS treatment decisions and generating hypotheses for future validation through pragmatic trials. Taken together, the study framework establishes a foundation for comparing other MS therapies and potentially extending clinical investigations to other chronic conditions.

### Limitations

Several limitations warrant consideration. First, although our imputation models for RRMS subtype and disability status performed well in held-out test sets, residual misclassification may still occur and could bias treatment effect estimates. To mitigate this, we calibrated imputed RRMS subtype against gold-standard registry labels, ensuring that all registry-confirmed RRMS patients were retained in the target population. Assuming modest misclassification in the non-registry EHR population, any residual error is likely to increase variance rather than introduce substantial bias. For disability outcome imputation, we incorporated calibration regression within the causal analysis framework to further reduce misclassification and improve validity of treatment effect estimates.

Second, unmeasured confounding remains an inherent challenge in observational research. Despite adjusting for a comprehensive set of codified and narrative EHR variables, residual differences, such as disease severity or socioeconomic factors, may persist between treatment groups. Our imputation approach enabled adjustment for unmeasured baseline disability status. Similar strategy could extend to other partially observed measures available through linked registry data. While *individual* socioeconomic factors are often difficult to capture in EHR data, future studies could incorporate *neighborhood-level* socioeconomic indicators based on residence and census data.

Third, because OCR was approved more recently than NTZ, temporal shifts in clinical practice and treatment effect are possible. To minimize this impact, we narrowed the study window to 2012-2020. Future studies could further address by adjusting for or matching on calendar year of treatment initiation.

## Conclusion

This study presents a robust framework for comparing treatment effects on disability outcomes across MS therapies that lack head-to-head RCT evidence. It offers a general roadmap for conducting scalable, bias-mitigated comparative effectiveness research using real-world clinical data. The semi-supervised doubly robust approach, combining machine learning-based imputation with a critical calibration step, addresses incomplete information that has previously limited the effective use of EHR data for causal inference. As a clinically relevant demonstration to address knowledge gaps, we provide real-world evidence that OCR is associated with lower two-year risk of moderate or severe disability compared with NTZ in RRMS by expanding the target population and applying bias-mitigated causal inference methods.

## Data Availability

Code for analysis and figures is available at <https://github.com/xialab2016/MS_OCRvsNTZ.git>.

We will publicly disseminate anonymous summary-level registry data and EHR data. The rationale for not sharing patient-level data is that patient-level clinical data (either de-identified information or limited protected health information containing dates of clinical events or even if anonymous due to concern for re-identification) are universally subject to the rules and regulation of each healthcare system, which may only be affiliated with but are not the same as the primary academic institutions of the study investigators. Sharing of de-identified EHR data with qualified external researchers by each of the study performance site may be permissible only after the approval of the respective Institutional Review Boards (IRBs), regulatory oversight agents of the healthcare systems (that own the clinical data) as well as the appropriate Data Usage Agreements (DUA) between institutions.

https://github.com/xialab2016/MS_OCRvsNTZ.git

## ACKNOWLEDGEMENT

### Data Access and Code Availability

Code for analysis and figures is available at <https://github.com/xialab2016/MS_OCRvsNTZ.git>. We will publicly disseminate anonymous summary-level registry data and EHR data. The rationale for not sharing patient-level data is that patient-level clinical data (either de-identified information or limited protected health information containing dates of clinical events or even if anonymous due to concern for re-identification) are universally subject to the rules and regulation of each healthcare system, which may only be affiliated with but are not the same as the primary academic institutions of the study investigators. Sharing of de-identified EHR data with qualified external researchers by each of the study performance site may be permissible only after the approval of the respective Institutional Review Boards (IRBs), regulatory oversight agents of the healthcare systems (that own the clinical data) as well as the appropriate Data Usage Agreements (DUA) between institutions.

