## Supplemental materials for "Ocrelizumab versus Natalizumab in Relapsing-Remitting Multiple Sclerosis: A Registry-Linked Electronic Health Records Study"

### SUPPLEMENTARY MATERIALS

#### S1. Supplementary Text

##### S1.1. Baseline Binary PDDS Imputation Method and Result

Baseline disability status is a potentially important confounder in causal analyses but frequently missing in real-world data. In addition to the clinician-rated disability measure based on the Expanded Disability Status Scale (EDSS) score, patient-reported disability measure based on the Patient Determined Disease Steps (PDDS) score provides crucial patient perspectives and complements EDSS. Because EDSS and PDDS are not interchangeable, inclusion of both disability measures at baseline for confounder adjustment more robustly mitigates potential bias. Thus, we imputed baseline disability status based on both EDSS (see **Main Methods – Clinician-Rated Disability Imputation**) and PDDS (see below).

PDDS is a validated patient-reported outcome measure that ranges from 0 to 8 in increments of 1. PDDS score of 4 indicates severe disability requiring full ambulatory support. We classified PDDS score ≥4 as severe disability and <4 as no, mild, or moderate disability. We computed the median PDDS score and defined the binary threshold (observed PDDS ≥4 vs <4) within consecutive six-month windows as the gold-standard patient-reported disability labels.

To impute the binary PDDS threshold (*i.e.,* imputed PDDS), we used the same features (see Main Methods - Feature Set, **Supplementary Table 1**) aggregated over consecutive 6-month windows and applied the same modeling framework as for EDSS imputation (see **Main Methods – Clinician-Rated Disability Imputation**). Specifically, we used an ensemble modeling approach combining standard supervised learning methods (*i.e.,* LASSO, XGBoost, Random Forest) with the semi-supervised LATTE method. We trained models using data from a randomly selected subset of 500 patients with available PDDS scores. Predicted likelihood (*i.e.,* probability) for severe patient-reported disability was derived using a cross-validated ensemble of all models (*i.e.,* "Ensemble" method).

Imputation model performance was evaluated in the held-out set comprising registry patients excluded from training by comparing predicted likelihood of severe disability to the gold-standard PDDS≥4 labels. For the binary PDDS classification, we reported AUC*,* sensitivity, specificity, and PPV.

For imputing the patient-reported binary disability status (*i.e.,* likelihood of PDDS≥4) during pre-treatment baseline, LATTE achieved an AUC of 0.879 in the held-out test set, while the Ensemble method slightly improved the prediction to AUC of 0.899 (**Supplementary Table 2**). Using a representative calibrated threshold of 0.95 for the likelihood of PDDS≥4 (to reach PPV≥0.90) for reporting and compared against observed labels, the ensemble model, which achieved a sensitivity of 0.081, specificity of 0.999, and PPV of 0.923 (at the representative threshold), was used for defining baseline patient-reported disability (**Supplementary Table 7**). Unlike the RRMS imputation to establish a cohort with the RRMS subtype, disability imputation (likelihood value ranging between 0 and 1) did not require a cut-off calibration threshold.

#### S1.2. Exploratory Analysis: NTZ versus BCD with RTX Projected onto Target Population

##### S1.2.1. *Heterogeneity of Patients Receiving RTX vs Patients Receiving OCR or NTZ*

Ocrelizumab (OCR) is part of the B-cell depletion (BCD) mechanism class, which also includes rituximab (RTX), ofatumumab (OFA) and ublituximab (UTX). During the study period (2012-2020, to ensure sufficient pre-treatment and post-treatment follow-up), the MGB EHR data contained no patients receiving OFA or UTX. RTX has been used as an off-label treatment of MS.

In the study dataset, RTX-treated and OCR-treated patients differed in their clinical and demographic characteristics (**Supplementary Table 3**). Compared with OCR-treated patients, RTX-treated patients had older age at disease-modifying therapy (DMT) initiation (mean 43.4 vs 42.2 years, $p<0.001$), a longer disease duration (6.6 vs 5.6 years, $p<0.001$), greater pre-treatment likelihood of EDSS ≥4 (0.25 vs 0.17, $p<0.001$), greater pre-treatment likelihood of PDDS≥4 (0.14 vs 0.13, $p<0.001$) and higher healthcare utilization.

##### S1.2.2. *Exploratory Causal Analysis Cohort Construction and Patient Profile: Method and Results*

To enable a fair comparison with natalizumab (NTZ), we projected the cohort to identify RTX-treated patients whose clinical profiles resemble those of the OCR- or NTZ-treated population in the primary causal analysis (**Supplementary Table 1**). To illustrate this projection, we displayed the distribution of RTX-treated patients relative to OCR- and NTZ-treated patients using a combined Partial Least Squares (PLS) regression and Uniform Manifold Approximation and Projection (UMAP) approaches for dimensionality reduction (**Supplementary Figure 3**).

Patient eligibility and treatment exposure criteria were consistent with those applied in the primary analysis (**Main Method – DMT Choice and Target Population**).

Among 1,057 eligible patients, we identified 196 patients with imputed and registry-derived RRMS subtype who had received RTX. For the exploratory causal analysis, only 66 RTX-treated patients had covariate profiles comparable to those in the primary causal analysis cohort (748 total: 543 OCR vs 205 NTZ) based on projected similarity in the multivariate covariate space (**Supplementary Figure 1**). We then pooled those 66 RTX-treated patients with OCR-treated patients to form the BCD treatment exposure group, yielding a final exploratory causal analysis cohort of 814 RRMS patients (BCD: 609; NTZ: 205).

Before covariate adjustment, treatment groups differed in baseline characteristics (**Supplementary Table 4**). Compared with BCD-treated RRMS patients at baseline, NTZ-treated patients had higher predicted pre-treatment disability measured by EDSS (OCR: 0.21 vs NTZ: 0.18; $p$=0.028), lower healthcare usage measured by EHR code counts (log-counts OCR: 2.40 vs NTZ: 2.71; $p$=0.001), and fewer MS diagnosis codes (log-counts OCR 2.03 vs NTZ: 2.38; $p$<0.001) These baseline differences suggest potential confounding by disease severity and healthcare utilization. After covariate adjustment by IPW, treatment groups were balanced on baseline characteristics (**Supplementary Figure 2**).

##### S1.2.3. *Exploratory Causal Analysis: Method and Results*

We applied the same doubly robust causal inference framework, combining propensity score modeling with calibration regression on imputed outcomes to minimize bias (see **Main Methods – Statistical Analysis for Estimating Average Treatment Effect**).

In the exploratory causal analysis using 814 patients with imputed RRMS subtype and imputed disability outcome (*i.e.,* likelihood of EDSS≥4.0 during two years after treatment initiation), BCD-treated patients (8.68%, 95% CI: 6.44%–10.93%) had a lower pEDSS, imputed two-year risk of moderate to severe disability, compared with NTZ-treated patients (14.53%, 95% CI: 10.60%–18.45%). The absolute risk difference was –5.84% (95% CI: –10.59 to –1.10; *p*=0.016), favoring BCD therapy (**Supplementary Figure 3**). Given the relatively modest sample size of the NTZ group, its ATE estimate had a wide 95% confidence interval and was less stable.

When analyzing the 647 patients with a registry-derived RRMS subtype, BCD (8.30%, 95% CI: 6.09%–10.51%) was associated with a lower pEDSS, imputed two-year risk of moderate to severe disability, compared with NTZ (14.24%, 95% CI, 10.03%–18.45%). The absolute difference was –5.94% (95% CI: –11.26 to –0.63; *p*=0.028), favoring BCD.

When analyzing the 351 patients with both a registry-derived RRMS subtype and observed EDSS outcomes (*i.e.,* gold-standard observed labels only), BCD (8.80%, 95% CI: 6.37%–11.23%) also showed a lower pEDSS, two-year disability risk of moderate to severe disability, compared with NTZ (14.49%, 95% CI: 9.00%–19.98%). The risk difference was –5.69% (95% CI: –12.41% to 1.04%; *p*=0.097). This final sensitivity analysis demonstrated a consistent trend, though it was not statistically significant, likely due to wider confidence intervals reflecting the smaller sample size and greater variability.

#### S2. Supplementary Figures

**Supplementary Figure 1. Patient profile projection in multivariate space.** Projection of patients receiving RTX onto the combined target patient population receiving OCR or NTZ. Patient embeddings were generated using a two-step dimensionality reduction approach that combined Partial Least Squares (PLS) regression and Uniform Manifold Approximation and Projection (UMAP). While OCR- and NTZ-treated patients (cyan and green) largely overlapped, most RTX-treated patients (red) formed a distinct cluster, with notable outliers, *i.e.,* RTX-outliers (purple). This pattern indicates that most RTX-treated patients generally did not share the same baseline profile as those in the primary causal analysis comparing OCR with NTZ.

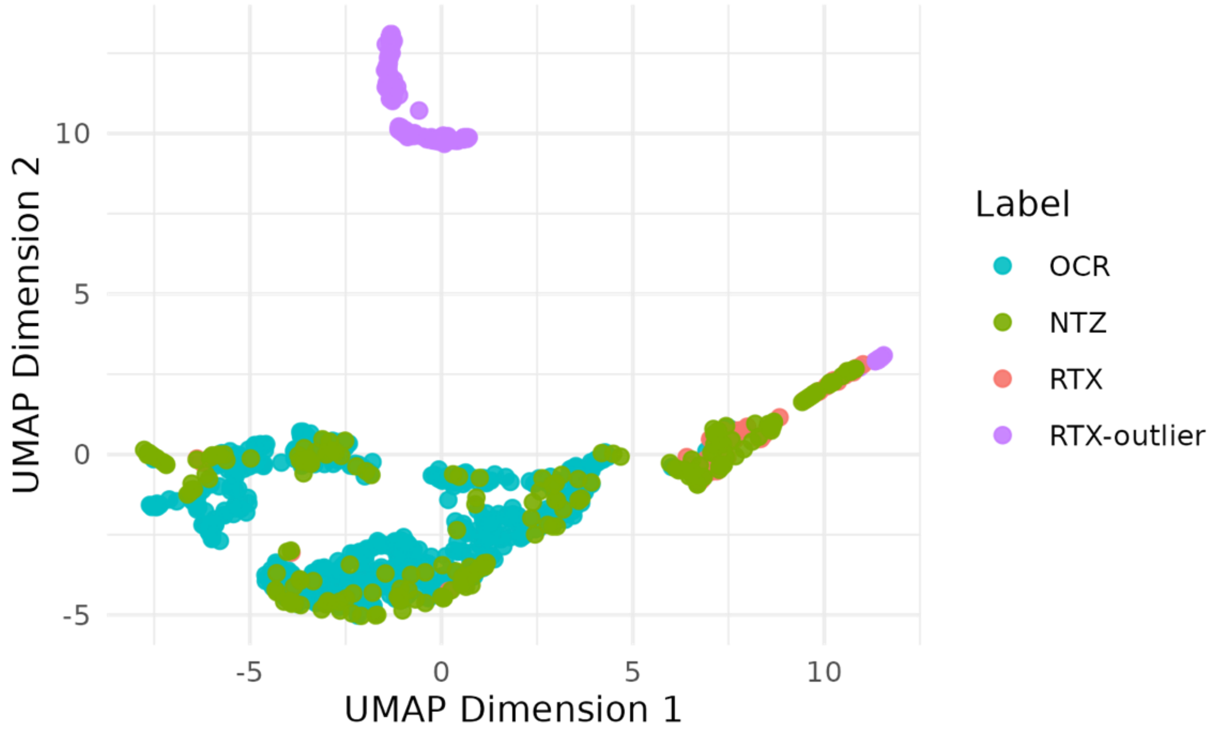

**Supplementary Figure 2. Covariate balancing.** P-values before and after inverse probability weighting (IPW) are shown for all covariates with non-zero coefficients in either the propensity score or outcome regression models in the **exploratory causal analysis** comparing the 2-year disability between **BCD** versus **NTZ**. The vertical dashed line at $p$=0.05 indicates the threshold for nominal significance. P-values were derived from chi-squared tests for categorical variables or two-sample Kolmogorov–Smirnov tests for continuous variables. All covariates were marginally balanced ($p$>0.05) post-weighting.

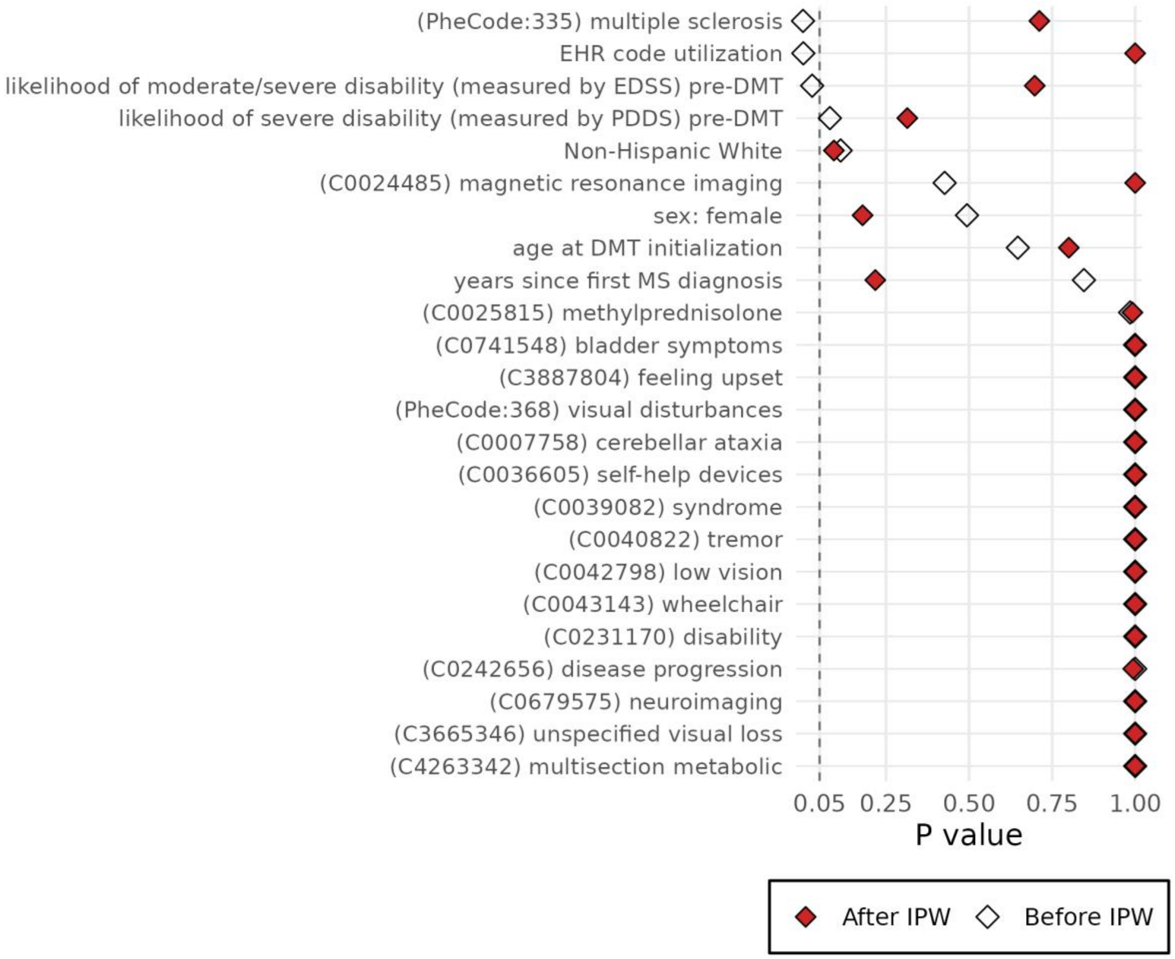

**Supplementary Figure 3. Average treatment effects of BCD vs NTZ**. Estimated average treatment effects (markers) on pEDSS, imputed likelihood of moderate or severe disability (EDSS≥4.0), during the two years after treatment initiation are shown with pointwise 95% confidence intervals (vertical bars). **BCD therapy (OCR or RTX)** was associated with a lower two-year risk of moderate or severe disability across analyses, when using pEDSS in patients with imputed RRMS subtype, imputed EDSS in registry patients with observed RRMS subtype or observed EDSS in registry patients with observed RRMS subtype. While point estimates remained similar, the confidence intervals widened when using registry-derived subtype and disability status.

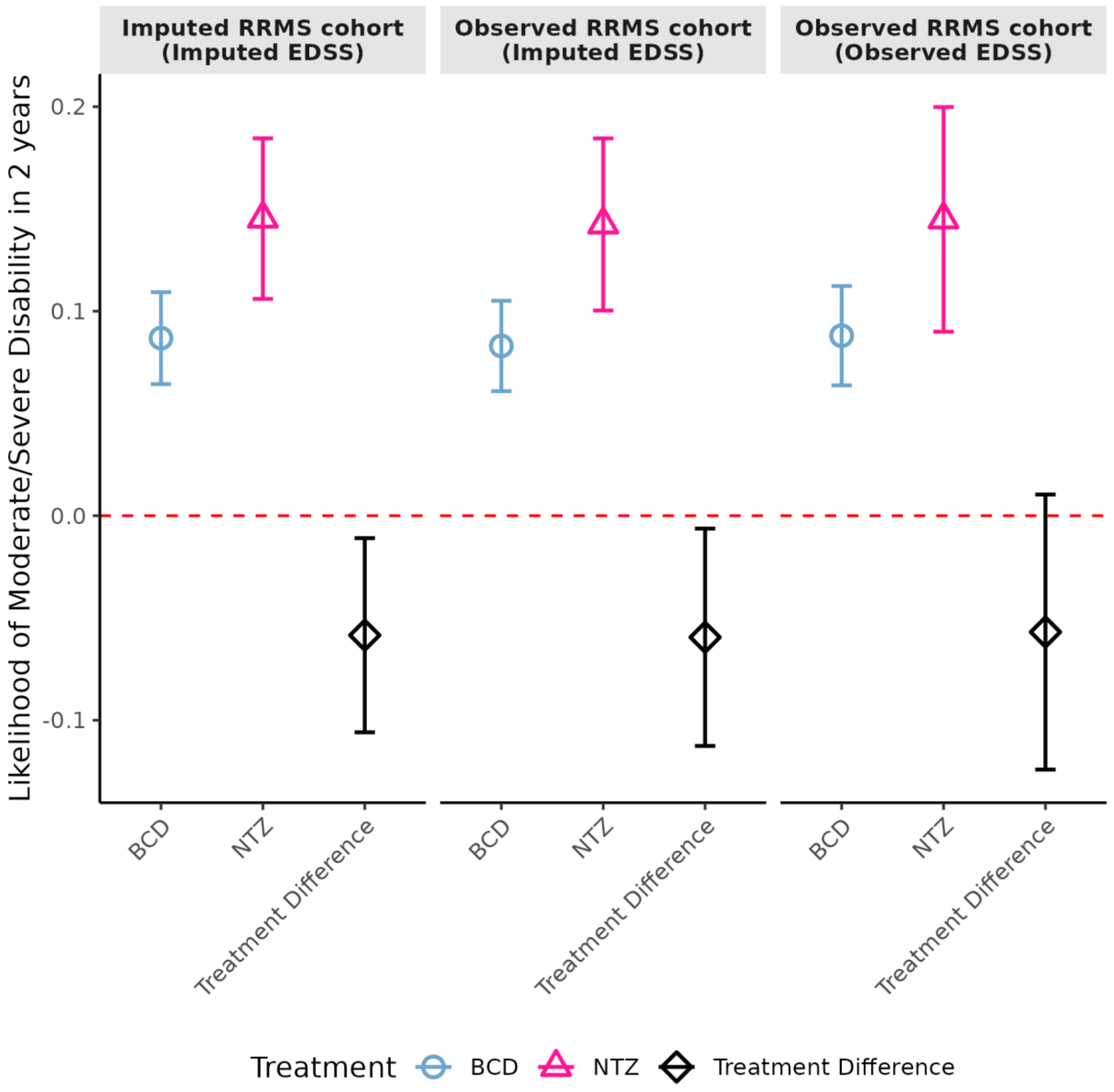

#### S3. Supplementary Tables

**Supplementary Table 1. Covariate balance assessment for the *primary* causal analysis** of comparing pEDSS, the two-year likelihood of moderate or severe disability, between OCR versus NTZ. Features with non-zero coefficients in either the propensity score or outcome regression models are shown. Feature values are shown as mean (SD) or patient number (% total). Codified EHR feature counts and NLP-extracted narrative concept counts were log-transformed. Univariate associations were tested using either chi-squared tests for binary covariates or two-sample Kolmogorov–Smirnov tests for continuous covariates (unweighted and weighted by inverse propensity scores). All covariates were marginally balanced ($p$>0.05) post-weighting.

| | | **Observed** | | | **Inverse Probability Weighted** | | |
|---|---|---|---|---|---|---|---|
| **Feature** | Overall | *OCR* | *NTZ* | *p-value* | *OCR* | *NTZ* | *p-value* |
| sex: female | 573 (77 %) | 413 (76 %) | 160 (78 %) | 0.566 | 425 (78 %) | 163 (80 %) | 0.658 |
| non-Hispanic White | 631 (84 %) | 464 (85 %) | 167 (81 %) | 0.180 | 460 (85 %) | 181 (88 %) | 0.174 |
| age at DMT initialization: years | 42.22 (11.53) | 42.87 (11.76) | 40.49 (10.76) | 0.755 | 42.47 (11.69) | 41.08 (10.55) | 0.994 |
| years since first MS diagnosis | 5.56 (5.31) | 5.73 (5.42) | 5.10 (4.97) | 0.739 | 5.36 (5.28) | 4.93 (5.11) | 1.000 |
| likelihood of moderate/severe disability (measured by EDSS) pre-DMT | 0.17 (0.18) | 0.16 (0.16) | 0.21 (0.20) | 0.003 | 0.17 (0.16) | 0.19 (0.21) | 0.979 |
| likelihood of severe disability (measured by PDDS) pre-DMT | 0.13 (0.07) | 0.12 (0.07) | 0.14 (0.08) | 0.020 | 0.13 (0.08) | 0.14 (0.13) | 1.000 |
| EHR code utilization | 2.64 (0.85) | 2.74 (0.76) | 2.40 (0.99) | <0.001 | 2.61 (0.88) | 2.77 (1.00) | 0.846 |
| (PheCode:335) multiple sclerosis | 2.31 (0.71) | 2.41 (0.59) | 2.03 (0.89) | <0.001 | 2.27 (0.71) | 2.41 (0.87) | 0.151 |
| (PheCode:350) abnormal movement | 0.17 (0.49) | 0.17 (0.50) | 0.16 (0.45) | 1.000 | 0.18 (0.50) | 0.22 (0.52) | 1.000 |

| | | | | | | | |
|---|---|---|---|---|---|---|---|
| (PheCode:368) visual disturbances | 0.10 (0.34) | 0.11 (0.34) | 0.10 (0.34) | 1.000 | 0.11 (0.34) | 0.08 (0.30) | 1.000 |
| (RXNORM:214582) glatiramer | 0.24 (0.53) | 0.20 (0.49) | 0.32 (0.61) | 0.998 | 0.18 (0.46) | 0.32 (0.67) | 1.000 |
| (C0000768) congenital abnormality | 0.72 (0.74) | 0.76 (0.74) | 0.62 (0.74) | 0.177 | 0.73 (0.80) | 0.79 (0.80) | 1.000 |
| (C0001807) aggressive behavior | 0.09 (0.30) | 0.07 (0.27) | 0.14 (0.36) | 0.994 | 0.10 (0.34) | 0.16 (0.36) | 1.000 |
| (C0004134) ataxia | 0.10 (0.35) | 0.09 (0.32) | 0.13 (0.42) | 1.000 | 0.08 (0.30) | 0.09 (0.34) | 1.000 |
| (C0004364) autoimmune diseases | 0.09 (0.32) | 0.10 (0.34) | 0.07 (0.27) | 1.000 | 0.09 (0.32) | 0.06 (0.23) | 1.000 |
| (C0009450) communicable diseases | 0.63 (0.70) | 0.73 (0.70) | 0.36 (0.62) | <0.001 | 0.71 (0.78) | 0.56 (0.74) | 0.862 |
| (C0012569) diplopia | 0.38 (0.65) | 0.37 (0.64) | 0.42 (0.68) | 1.000 | 0.36 (0.62) | 0.56 (0.84) | 1.000 |
| (C0013362) dysarthria | 0.08 (0.29) | 0.08 (0.27) | 0.10 (0.33) | 1.000 | 0.08 (0.27) | 0.05 (0.24) | 1.000 |
| (C0013987) emotions | 0.18 (0.44) | 0.17 (0.44) | 0.18 (0.43) | 1.000 | 0.16 (0.42) | 0.21 (0.44) | 1.000 |
| (C0015726) fear (mental process) | 0.32 (0.55) | 0.37 (0.57) | 0.20 (0.48) | 0.022 | 0.35 (0.57) | 0.55 (0.79) | 0.775 |
| (C0018786) hearing tests | 0.73 (0.71) | 0.78 (0.72) | 0.59 (0.69) | 0.102 | 0.72 (0.71) | 0.76 (0.76) | 1.000 |
| (C0023185) learning | 0.15 (0.43) | 0.13 (0.40) | 0.21 (0.51) | 1.000 | 0.12 (0.40) | 0.26 (0.60) | 1.000 |
| (C0024485) magnetic resonance imaging | 0.13 (0.34) | 0.11 (0.31) | 0.21 (0.39) | 0.417 | 0.12 (0.32) | 0.15 (0.33) | 1.000 |
| (C0025815) methylprednisolone | 0.42 (0.69) | 0.38 (0.62) | 0.54 (0.83) | 0.977 | 0.35 (0.60) | 0.60 (0.84) | 0.854 |
| (C0036429) sclerosis | 0.45 (0.64) | 0.49 (0.68) | 0.33 (0.54) | 0.774 | 0.46 (0.65) | 0.58 (0.77) | 1.000 |
| (C0036605) self-help devices | 0.05 (0.22) | 0.05 (0.21) | 0.04 (0.23) | 1.000 | 0.05 (0.23) | 0.09 (0.37) | 1.000 |

| | | | | | | | |
|---|---|---|---|---|---|---|---|
| (C0038435) stress | 0.34 (0.63) | 0.36 (0.65) | 0.30 (0.57) | 1.000 | 0.33 (0.62) | 0.50 (0.71) | 0.904 |
| (C0042024) urinary incontinence | 0.16 (0.48) | 0.17 (0.51) | 0.14 (0.40) | 1.000 | 0.19 (0.57) | 0.18 (0.40) | 1.000 |
| (C0042798) low vision | 0.06 (0.27) | 0.06 (0.26) | 0.07 (0.27) | 1.000 | 0.05 (0.25) | 0.06 (0.30) | 1.000 |
| (C0043143) wheelchair | 0.06 (0.30) | 0.06 (0.29) | 0.08 (0.34) | 1.000 | 0.08 (0.35) | 0.14 (0.52) | 1.000 |
| (C0080274) urinary retention | 0.28 (0.59) | 0.27 (0.58) | 0.29 (0.60) | 1.000 | 0.28 (0.60) | 0.54 (0.90) | 0.704 |
| (C0221198) lesion | 1.53 (0.76) | 1.61 (0.72) | 1.32 (0.84) | 0.017 | 1.52 (0.81) | 1.63 (0.82) | 0.557 |
| (C0231170) disability | 0.27 (0.59) | 0.26 (0.60) | 0.28 (0.56) | 1.000 | 0.25 (0.58) | 0.30 (0.60) | 1.000 |
| (C0338656) impaired cognition | 0.14 (0.42) | 0.12 (0.37) | 0.20 (0.52) | 1.000 | 0.12 (0.41) | 0.24 (0.58) | 1.000 |
| (C0358514) diagnostic agents | 0.25 (0.48) | 0.27 (0.50) | 0.21 (0.43) | 1.000 | 0.31 (0.64) | 0.38 (0.53) | 0.989 |
| (C0439044) living alone | 0.15 (0.46) | 0.15 (0.45) | 0.16 (0.46) | 1.000 | 0.15 (0.44) | 0.19 (0.44) | 1.000 |
| (C0456909) blindness | 0.10 (0.34) | 0.11 (0.35) | 0.09 (0.31) | 1.000 | 0.13 (0.37) | 0.12 (0.37) | 1.000 |
| (C0600116) does speak | 0.24 (0.48) | 0.23 (0.48) | 0.26 (0.47) | 1.000 | 0.24 (0.49) | 0.44 (0.67) | 0.796 |
| (C0679575) neuroimaging | 0.14 (0.36) | 0.13 (0.35) | 0.14 (0.38) | 1.000 | 0.12 (0.34) | 0.11 (0.31) | 1.000 |
| (C0683525) treatment options | 0.50 (0.60) | 0.51 (0.59) | 0.50 (0.64) | 1.000 | 0.47 (0.59) | 0.65 (0.71) | 0.982 |
| (C0741548) bladder symptoms | 0.11 (0.34) | 0.09 (0.33) | 0.16 (0.36) | 0.973 | 0.08 (0.32) | 0.10 (0.28) | 1.000 |
| (C3714552) weakness | 0.82 (0.85) | 0.81 (0.85) | 0.84 (0.86) | 1.000 | 0.77 (0.86) | 1.02 (0.96) | 0.579 |
| (C3887804) feeling upset | 0.09 (0.31) | 0.07 (0.29) | 0.13 (0.35) | 1.000 | 0.08 (0.30) | 0.13 (0.34) | 1.000 |
| (C4263342) multisection metabolic | 0.08 (0.25) | 0.07 (0.23) | 0.08 (0.27) | 1.000 | 0.08 (0.25) | 0.17 (0.41) | 1.000 |

| | | | | | | | |
|---|---|---|---|---|---|---|---|
| (C5203119) intensity and distress | 0.55 (0.77) | 0.57 (0.79) | 0.49 (0.70) | 1.000 | 0.57 (0.80) | 0.64 (0.94) | 1.000 |

**Supplementary Table 2. Imputation predictive performance.** Area under the receiver operating characteristic curve (AUC) with 95% confidence intervals for imputing patient-reported severe disability (**PDDS ≥ 4**) by the different machine learning methods is shown.

| **Imputed Outcome** | Ensemble Method * | LATTE | LASSO | Random Forest | XGBoost | **Training Set**: patient number (total records) | **Test Set**: patient number (total records) |
|---|---|---|---|---|---|---|---|
| PDDS≥4 | 0.899 (0.874-0.924) | 0.879 (0.852-0.907) | 0.867 (0.840-0.894) | 0.754 (0.700-0.807) | 0.828 (0.791-0.864) | 500 (774) | 1,982 (3,205) |

**Supplementary Table 3. Heterogeneity of Patients Receiving RTX vs Patients Receiving OCR or NTZ.** Demographic and standard clinical characteristics of RTX-treated patients were compared with OCR- or NTZ-treated patients from the primary causal analysis (*i.e.*, the target population). Feature values are shown as mean (SD) or patient number (% total). P-values reflect statistical comparisons between groups using either chi-squared tests for binary covariates or two-sample Kolmogorov–Smirnov tests for continuous covariates.

| | Observed | | |
|---|---|---|---|
| Variable | Target population (OCR+NTZ) | RTX | p-value |
| sex: female | 573 (77 %) | 148 (76 %) | 0.748 |
| Non-Hispanic White | 631 (84 %) | 155 (79 %) | 0.078 |
| age at DMT initialization | 42.22 (11.53) | 43.41 (11.32) | <0.001 |
| years since first MS diagnosis | 5.56 (5.31) | 6.59 (5.30) | <0.001 |
| likelihood of moderate/severe disability (measured by EDSS) pre-DMT | 0.17 (0.18) | 0.25 (0.24) | <0.001 |
| likelihood of severe disability (measured by PDDS) pre-DMT | 0.13 (0.07) | 0.14 (0.08) | <0.001 |
| EHR code utilization | 2.64 (0.85) | 2.74 (0.85) | <0.001 |
| (PheCode:335) multiple sclerosis | 2.31 (0.71) | 2.33 (0.73) | <0.001 |

**Supplementary Table 4. Covariate balance assessment for the *exploratory* causal analysis** of comparing the 2-year disability between BCD versus NTZ. Features with non-zero coefficients in either the propensity score or outcome regression models are shown. Feature values are shown as mean (SD) or patient number (% total). Codified EHR feature counts and NLP-extracted narrative concept counts were log-transformed. Univariate associations were tested using either chi-squared tests for binary covariates or two-sample Kolmogorov–Smirnov tests for continuous covariates (unweighted and weighted by inverse propensity scores). All covariates were marginally balanced (p>0.05) post-weighting.

| | | **Observed** | | | **Inverse Probability Weighted** | | |
|---|---|---|---|---|---|---|---|
| **Feature** | Overall | *OCR* | *NTZ* | *p-value* | *OCR* | *NTZ* | *p-value* |
| sex: female | 621 (76 %) | 461 (76 %) | 160 (78 %) | 0.494 | 459 (75 %) | 163 (80 %) | 0.179 |
| non Hispanic White | 691 (85 %) | 524 (86 %) | 167 (81 %) | 0.113 | 510 (84 %) | 181 (88 %) | 0.093 |
| age at DMT initialization | 42.37 (11.55) | 43.00 (11.74) | 40.49 (10.76) | 0.647 | 42.18 (11.96) | 41.08 (10.55) | 0.800 |
| years since first MS diagnosis | 5.61 (5.34) | 5.78 (5.45) | 5.10 (4.97) | 0.846 | 6.17 (5.33) | 4.93 (5.11) | 0.218 |
| likelihood of moderate/severe disability (measured by EDSS) pre-DMT | 0.18 (0.19) | 0.18 (0.19) | 0.21 (0.20) | 0.028 | 0.24 (0.23) | 0.19 (0.21) | 0.698 |
| likelihood of severe disability (measured by PDDS) pre-DMT | 0.13 (0.07) | 0.13 (0.07) | 0.14 (0.08) | 0.081 | 0.15 (0.08) | 0.14 (0.13) | 0.314 |
| EHR code utilization | 2.63 (0.85) | 2.71 (0.78) | 2.40 (0.99) | 0.001 | 2.55 (0.91) | 2.77 (1.00) | 1.000 |
| (PheCode:335) multiple sclerosis | 2.29 (0.72) | 2.38 (0.63) | 2.03 (0.89) | <0.001 | 2.21 (0.77) | 2.41 (0.87) | 0.712 |
| (PheCode:368) visual disturbances | 0.10 (0.33) | 0.10 (0.33) | 0.10 (0.34) | 1.000 | 0.08 (0.27) | 0.08 (0.30) | 1.000 |
| (C0007758) cerebellar ataxia | 0.10 (0.32) | 0.08 (0.28) | 0.13 (0.41) | 1.000 | 0.15 (0.37) | 0.09 (0.33) | 1.000 |
| (C0024485) magnetic resonance imaging | 0.13 (0.34) | 0.11 (0.32) | 0.21 (0.39) | 0.426 | 0.13 (0.34) | 0.15 (0.33) | 1.000 |
| (C0025815) methylprednisolone | 0.43 (0.69) | 0.39 (0.64) | 0.54 (0.83) | 0.985 | 0.51 (0.73) | 0.60 (0.84) | 0.992 |
| (C0036605) self-help devices | 0.05 (0.23) | 0.05 (0.23) | 0.04 (0.23) | 1.000 | 0.07 (0.36) | 0.09 (0.37) | 1.000 |

| | | | | | | | |
|---|---|---|---|---|---|---|---|
| (C0039082) syndrome | 0.15 (0.44) | 0.16 (0.47) | 0.11 (0.33) | 1.000 | 0.10 (0.37) | 0.17 (0.40) | 1.000 |
| (C0040822) tremor | 0.25 (0.52) | 0.26 (0.53) | 0.24 (0.48) | 1.000 | 0.22 (0.50) | 0.30 (0.58) | 1.000 |
| (C0042798) low vision | 0.06 (0.27) | 0.06 (0.27) | 0.07 (0.27) | 1.000 | 0.09 (0.37) | 0.06 (0.30) | 1.000 |
| (C0043143) wheelchair | 0.07 (0.32) | 0.07 (0.31) | 0.08 (0.34) | 1.000 | 0.11 (0.37) | 0.14 (0.52) | 1.000 |
| (C0231170) disability | 0.27 (0.59) | 0.26 (0.60) | 0.28 (0.56) | 1.000 | 0.22 (0.56) | 0.30 (0.60) | 1.000 |
| (C0242656) disease progression | 0.13 (0.32) | 0.13 (0.30) | 0.15 (0.38) | 1.000 | 0.08 (0.26) | 0.22 (0.43) | 0.994 |
| (C0679575) neuroimaging | 0.13 (0.35) | 0.13 (0.34) | 0.14 (0.38) | 1.000 | 0.10 (0.29) | 0.11 (0.31) | 1.000 |
| (C0741548) bladder symptoms | 0.11 (0.35) | 0.10 (0.35) | 0.16 (0.36) | 1.000 | 0.14 (0.36) | 0.10 (0.28) | 1.000 |
| (C3665346) unspecified visual loss | 0.10 (0.36) | 0.10 (0.38) | 0.08 (0.30) | 1.000 | 0.06 (0.29) | 0.08 (0.32) | 1.000 |
| (C3887804) feeling upset | 0.08 (0.30) | 0.07 (0.28) | 0.13 (0.35) | 1.000 | 0.07 (0.26) | 0.13 (0.34) | 1.000 |
| (C4263342) multisection metabolic | 0.07 (0.24) | 0.07 (0.23) | 0.08 (0.27) | 1.000 | 0.09 (0.24) | 0.17 (0.41) | 1.000 |

**Supplementary Table 5. Imputation predictive performance:** Sensitivity, positive predictive value (PPV) and negative predictive value (NPV) for imputing the **RRMS** subtype by the different machine learning methods using a *pre-specified* calibrated threshold of ≥0.63 (to reach PPV≥0.90 for the **Ensemble** method) is shown.

| **Method \ Metric** | Specificity | Sensitivity | PPV | NPV | Calibration Method |
|---|---|---|---|---|---|
| LASSO | 0.849 | 0.698 | 0.898 | 0.597 | 0.63 |
| Random Forest | 0.794 | 0.740 | 0.872 | 0.616 | 0.63 |
| XGBoost | 0.760 | 0.787 | 0.861 | 0.652 | 0.63 |
| LATTE | 0.874 | 0.696 | 0.913 | 0.602 | 0.63 |
| Ensemble | 0.831 | 0.808 | 0.901 | 0.695 | 0.63 |

**Supplementary Table 6. Imputation predictive performance:** sensitivity, positive predictive value (PPV) and negative predictive value (NPV) for imputing pEDSS, clinician-rated moderate to severe disability (**EDSS≥4.0**), using a *representative* threshold of 0.5 (for reporting performance metrics) by the different machine learning methods or using a calibrated threshold of ≥0.78 (to reach PPV≥0.90 for the **Ensemble** method) is shown. Unlike the RRMS imputation to establish a cohort with the RRMS subtype, disability imputation (likelihood value ranging between 0 and 1) did not require a cut-off calibration threshold. We applied calibration only for the final ensemble method, while individual machine learning method was not calibrated. Of note, since the **predicted likelihood of EDDS ≥4.0** using LATTE are all smaller than 0.78, some performance metrics such as specificity, PPV or NPV may be NA.

| **Method \ Metric** | Specificity | Sensitivity | PPV | NPV | Calibration Threshold |
|---|---|---|---|---|---|
| LASSO | 0.961 | 0.509 | 0.806 | 0.860 | 0.5 |
| Random Forest | 0.981 | 0.412 | 0.874 | 0.840 | 0.5 |
| XGBoost | 0.945 | 0.504 | 0.746 | 0.857 | 0.5 |
| LATTE | NA | 1.000 | 0.242 | NA | 0.5 |
| Ensemble | 0.956 | 0.670 | 0.828 | 0.901 | 0.5 |

| **Method \ Metric** | Specificity | Sensitivity | PPV | NPV | Calibration Threshold |
|---|---|---|---|---|---|
| LASSO | 0.986 | 0.322 | 0.879 | 0.820 | 0.78 |
| Random Forest | 0.999 | 0.061 | 0.965 | 0.770 | 0.78 |
| XGBoost | 0.966 | 0.424 | 0.800 | 0.840 | 0.78 |
| LATTE | 1.000 | 0.000 | NA | 0.758 | 0.78 |
| Ensemble | 0.983 | 0.506 | 0.902 | 0.862 | 0.78 |

**Supplementary Table 7. Imputation predictive performance**: Sensitivity, positive predictive value (PPV) and negative predictive value (NPV) for imputing patient-reported severe disability (**PDDS ≥ 4**) using a *representative* calibration threshold of 0.5 (for reporting performance metrics) by the different machine learning methods or using a calibrated threshold of ≥0.95 (to reach PPV≥0.90 for the **Ensemble** method) is shown. Unlike the RRMS imputation to establish a cohort with the RRMS subtype, disability imputation (likelihood value ranging between 0 and 1) did not require a cut-off calibration threshold. We applied calibration only for the final ensemble method, while individual machine learning method was not calibrated. Of note, since the **predicted likelihood of PDDS ≥4** using random forest and LATTE are smaller than 0.95, some performance metrics such as specificity, PPV, or NPV may be NA.

| **Method \ Metric** | Specificity | Sensitivity | PPV | NPV | Calibration Threshold |
|---|---|---|---|---|---|
| LASSO | 0.951 | 0.372 | 0.579 | 0.893 | 0.5 |
| Random Forest | 0.977 | 0.318 | 0.712 | 0.888 | 0.5 |
| XGBoost | 0.916 | 0.486 | 0.511 | 0.908 | 0.5 |
| LATTE | NA | 1.000 | 0.153 | NA | 0.5 |
| Ensemble | 0.906 | 0.655 | 0.557 | 0.936 | 0.5 |

| **Method \ Metric** | Specificity | Sensitivity | PPV | NPV | Calibration Threshold |
|---|---|---|---|---|---|
| LASSO | 0.998 | 0.115 | 0.895 | 0.862 | 0.95 |
| Random Forest | 1.000 | 0.000 | NA | 0.847 | 0.95 |
| XGBoost | 0.973 | 0.243 | 0.621 | 0.877 | 0.95 |
| LATTE | 1.000 | 0.000 | NA | 0.847 | 0.95 |
| Ensemble | 0.999 | 0.081 | 0.923 | 0.857 | 0.95 |
